# The potential impact of *Anopheles stephensi* establishment on the transmission of *Plasmodium falciparum* in Ethiopia and prospective control measures

**DOI:** 10.1101/2021.08.19.21262272

**Authors:** Arran Hamlet, Dereje Dengela, J. Eric Tongren, Fitsum G Tadesse, Teun Bousema, Marianne Sinka, Aklilu Seyoum, Seth R. Irish, Jennifer S. Armistead, Thomas Churcher

**Affiliations:** MRC Centre for Global Infectious Disease Analysis; and the Abdul Latif Jameel Institute for Disease and Emergency Analytics, School of Public Health, Imperial College London, London, UK; PMI VectorLink Project, Abt Associates, 6130 Executive Blvd, Rockville, MD 20852, USA; US President’s Malaria Initiative (PMI), Addis Ababa, Ethiopia; Malaria Branch, Division of Parasitic Diseases and Malaria, Center for Global Health, Centers for Disease Control and Prevention, Atlanta, GA, USA; Armauer Hansen Research Institute, Addis Ababa, Ethiopia; Radboud Institute for Health Sciences, Radboud University Medical Center, Nijmegen, The Netherlands; Department of Immunology and Infection, London School of Hygiene and Tropical Medicine, London, UK; Department of Zoology, University of Oxford, Oxford, United Kingdom, OX1 3SZ; US President’s Malaria Initiative, Entomology Branch Division of Parasitic Diseases and Malaria, Center for Global Health, Centers for Disease Control and Prevention, Atlanta, GA, USA; U.S. President’s Malaria Initiative, U.S. Agency for International Development, Washington, D.C., USA

**Keywords:** Anopheles stephensi, malaria, mathematical modelling, Ethiopia, insecticide, invasive, vector control, Indoor residual spraying, Larval source management, Insecticide treated nets, invasive mosquito, Djibouti

## Abstract

**Background:** Sub-Saharan Africa has seen substantial reductions in cases and deaths due to malaria over the past two decades. While this reduction is primarily due to an increasing expansion of interventions, urbanisation has played its part as urban areas typically experience substantially less malaria transmission than rural areas. However, this may be partially lost with the invasion and establishment of *Anopheles stephensi. An. stephensi*, the primary urban malaria vector in Asia, was first detected in Africa during 2012 in Djibouti and was subsequently identified in Ethiopia in 2016, and later in Sudan and Somalia. In Djibouti, malaria cases have increased 30-fold from 2012 to 2019 though the impact in the wider region remains unclear.

**Methods:** Here we have adapted an existing model of mechanistic malaria transmission to estimate the increase in vector density required to explain the trends in malaria cases seen in Djibouti. To account for the observed plasticity in *An. stephensi* behaviour, and the unknowns of how it will establish in a novel environment, we sample behavioural parameters in order to account for a wide range of uncertainty. This quantification is then applied to Ethiopia, considering temperature-dependent extrinsic incubation periods, pre-existing vector-control interventions and *Plasmodium falciparum* prevalence in order to assess the potential impact of *An. stephensi* establishment on *P. falciparum* transmission. Following this, we estimate the potential impact of scaling up ITN (insecticide treated nets)/IRS (indoor residual spraying) and implementing piperonyl butoxide (PBO) ITNs and larval source management,, as well as their economic costs.

**Results:** We estimate that annual *P. falciparum* malaria cases could increase by 50% (95% CI 14-90) if no additional interventions are implemented. The implementation of sufficient control measures to reduce malaria transmission to pre-*stephensi* levels will cost hundreds of millions of USD.

**Conclusions:** Substantial heterogeneity across the country is predicted and large increases in vector control interventions could be needed to prevent a major public health emergency.

## Background

Sub-Saharan Africa, where 94% of the global malaria burden occurs, has seen a substantial reduction in cases and deaths due to malaria over the past two decades [1]. While this reduction is primarily due to an increase in investment and expansion of interventions such as insecticide treated nets (ITNs), indoor residual spraying (IRS), and diagnosis and treatment, urbanisation also has played a part.

Africa has experienced rapid urbanization in recent years, rising from 31.5% of the population living in urban areas in 1990, to 42.5% in 2018. By 2050, approximately 60% of the population is expected to live in urban areas [2]. Planned urbanisation will likely have a positive effect on reducing the malaria burden in Africa, as urban areas typically experience substantially lower rates of malaria transmission than rural areas [3]. Primarily, this is thought to be due to improved housing and the reduced availability of suitable larval habitats for African *Anopheles* vector species [4].

The protective effect conferred by urbanization may be partially lost with the invasion and establishment of *Anopheles stephensi. An. stephensi* is found throughout South Asia, where it is capable of transmitting both *Plasmodium falciparum* and *P. vivax* parasites [5] in a diverse set of habitats, from rural to highly urban settings [6]. The success of this vector in urban locations is due to its ability to utilise water tanks, wells, and other artificial containers as larval habitats [7, 8]. Furthermore, it has shown substantial resistance to water pollution [9]. In the last decade *Anopheles stephensi* has been discovered outside of its traditional endemic region in Asia and was first detected in Djibouti in 2012 [10].

Prior to 2013, Djibouti had reported less than 3000 cases of malaria per year. Following the year of initial detection of *An. stephensi*, cases have increased substantially, and in 2019 there were 49,402 confirmed cases of malaria [11]. While causation has not been established between increasing *An. stephensi* detection and malaria incidence in Djibouti, it has been heavily implicated [12]. *Anopheles stephensi* has now been found in Sudan, Somalia, and Ethiopia [13-17].

Here we quantify the potential impact of *An. stephensi* invasion in Djibouti on malaria transmission in order to make projections about what could happen in Ethiopia, where the species has been found at numerous sites and is spreading [18]. Translating the invasion of *An. stephensi* to its public health impact is difficult due to uncertainty in its vectorial capacity and how public health entities and governments will respond. Different illustrative scenarios are investigated, exploring the public health impact of different vector control interventions.

## Methods

### Mechanistic malaria model

A deterministic version of a well-established compartmental model of *P. falciparum* malaria transmission [19-24] was utilised. The human population was split into susceptible or infected individuals with those infected being either asymptomatic, having clinical disease, sub-microscopic infections, having been treated, or being in a period of prophylaxis following treatment. The numbers of mosquito larvae, pupae and adults were simulated with adult mosquitoes being either susceptible, exposed, or infectious (after the extrinsic incubation period, EIP). The model accounts for heterogeneity in transmission as well as age-dependent mosquito biting rates and the acquisition of natural immunity. The model is summarised in Additional File Section 1 [19, 20, 22, 23, 25-28].

### Vector bionomics and Latin hypercube sampling

The epidemiological impact of *An. stephensi* invasion will depend on the characteristics of the vector in the new environment. Vector behaviours such as crepuscular biting, and resting outside of houses could translate, compared to other African *Anopheles* species, to a reduced efficacy of interventions such as ITN and IRS, and therefore accurate parameterisation is important. A literature search was undertaken to find *An. stephensi*-specific bionomic data to parameterise the mechanistic model. These included the daily mortality, proportion of blood meals taken on humans, endophily (the proportion of time spent in human dwellings), bites taken indoors and in sleeping spaces. Data on *An. stephensi* behaviour within Africa was sparse, and so estimates were primarily acquired from studies in Iran, India and Pakistan, with limited data from Ethiopia. A full list of studies and parameter range estimates are in the Additional File Section 2 [5, 29-55]

Due to the relatively low number of studies from which the data were collected, and large uncertainty around how the vector would behave, we incorporated parameter sampling in the model fitting and extrapolation stage. This was done through taking the median value from the data, and sampling from values 25% smaller and 25% larger unless otherwise stated (Additional File Section 2). From this we undertook Latin hypercube sampling (LHS) which is a statistical method for generating near-random samples of parameter values from a multidimensional distribution. This allowed us to efficiently sample different parameter combinations in order to generate broad uncertainty in predictions [56].

The efficacy of current and future ITNs will also depend on the level of pyrethroid resistance. Substantial pyrethroid resistance has been found in Ethiopia, with a mean across years and sites of 57% mosquito survival upon exposure to pyrethroids in a discriminating dose bioassay [18]. This value was assumed throughout the country. There is no clear picture of how *An. stephensi* abundance will change seasonally in Ethiopia, and so mosquito density was assumed to remain consistent throughout the year, similar to recent patterns reported in Djibouti (Seyfarth et al 2019). Ethiopia is geographically diverse, with many regions at high altitudes, reducing malaria transmission. The relationship of the extrinsic incubation period and temperature were provided from Stopard et al. (2021) [33]. How this influences transmission is outlined in the Additional File Section 2 and Section 3.

### Fitting to Djibouti data

The severity of the public health impact of *An. stephensi* invasion will depend on the number of mosquitoes per person (vector density) and how it changes over time. In the absence of information on the speed and magnitude of *An. stephensi* establishment in Ethiopia, we made the simplest assumption that density increases in a sigmoidal manner that mirrors what has happened in Djibouti. Here, given the low levels of malaria reported prior to *An. stephensi* detection we assumed that the increase is singly due to *An. stephensi*, though native *Anopheles* species such as *An. arabiensis* are present. This process is conducted in conjunction with 200 LHS parameterisations of vector bionomics. From this, we take the 100 best LHS combinations as defined by their likelihood, and calculate the 2.5, 50 and 97.5% quantiles. This provides us with estimates of the vector density required to explain malaria incidence in Djibouti (Additional File Section 3). We then take the vector density required to explain the final year where it appears to have plateaued, 2019, and apply this to Ethiopia. Here the increase in vector density does not have an effect on pre-existing population dynamics, in the absence of existing information on the interaction of *An. stephensi* and local *Anopheles* vectors we do not model interaction or replacement, and so the existing vector density is increased by the value estimated above.

### Current level of disease

Estimates of the geographical distribution of malaria within Ethiopia were generated from malaria slide prevalence in 2–10-year-olds estimates extracted using the Malaria Atlas Project (MAP) R package, malariaAtlas [57]. These were aggregated to the 3rd administrative division (woreda) using a population weighted mean. Estimates were adjusted so that the number of cases predicted by the model matched those reported in the World Malaria Report (WMR). In 2019 (the last year data were available) a total of 738,155 cases of *P. falciparum* malaria were reported in Ethiopia. Here we concentrate on Falciparum malaria as this is the dominant malaria species with the biggest public health impact. Current estimates of the level of malaria control in Ethiopia are assumed to be maintained throughout the period of *An. stephensi* invasion. ITN usage was provided at the 1^st^ administrative division (region) assuming pyrethroid-only ITNs, and IRS coverage at the woreda level [58]. Human population sizes were taken from UN World Population Prospects [29]. Historical use of antimalarial drug treatment was also extracted from malariaAtlas (Additional File Section 2) and is assumed to remain constant over time. In order to reduce computational demands, prevalence was rounded to 0, 1, 5, 10, 15, 25%, historical ITN/IRS/treatment to the nearest 20%, and EIP the nearest 1 day. This reduced the number of administrative locations considered from 690 3^rd^ administrative (woreda) units to 64 administrative groupings (see Additional File Section 2).

### Projections of future malaria burden

Models were used to project the change in malaria prevalence over time following the invasion of *An. stephensi*. Here, we assumed an equal and simultaneous introduction in all groupings, which while unrealistic is the most parsimonious response (Additional File, Section 4 [28, 59-61]). Simulations were run for each administrative grouping. It is likely that *An. stephensi* will not invade all regions of the country [31], and that invasion in areas above an altitude of 2000m is unlikely to result in an increased malaria burden due to the low temperatures. We therefore estimated the increase in the number of malaria cases in the population within areas found suitable by previous research [31], and beneath 2000 meters [32].

Different vector control interventions are considered to mitigate the impact of *An. stephensi* invasion. As possible scenarios, we investigated the potential impact of increasing ITN usage, deploying IRS and larval source management (LSM). ITN usage was increased from its current level (Additional File Section 2) up to 80% usage assuming mass distribution every 3-years (assuming pyrethroid-only ITNs). For the implementation of synergist piperonyl butoxide (PBO) via PBO-pyrethroid ITNs, usage of 0% was assumed and scale-up involved replacing all existing standard pyrethroid-only ITNs. Pyrethroid-only and PBO-pyrethroid ITNs efficacy and its decay over their lifespan is estimated from a meta-analyses of experimental hut trial data as detailed in Churcher et al., (2016) [27]. Models parameterised with these data have been able to recreate the epidemiological impact of the mass distribution of pyrethroid-only ITNs, PBO-pyrethroid ITNs and IRS measured in cluster randomised control trials in Tanzania and Uganda [24]. In costing future interventions, we subtracted the cost of existing ITN usage, as we were calculating the additional cost. IRS covering 80% of structures of human habitation is conducted annually with a long-lasting product which the local mosquito population is fully susceptible to; this decays throughout the year as previously described [62]. The impact of LSM against *An. stephensi* is highly unclear but in the absence of this information it was assumed to be constant and at a level to reduce adult emergence across the year by 40%. These values were chosen based on pre-existing usage/coverage in Ethiopia (Additional File Section 2) and from discussions with implementation partners at the Presidents Malaria Initiative (PMI) centred around desired values and feasibility. Note that some of the intervention combinations are not currently recommended by the World Health Organisation (WHO) but are considered for completeness. To illustrate the broad economic cost of this additional vector control, simple estimates of total costs are provided. Approximate estimates of the cost of intervention (purchasing, delivering, and applying) were provided from literature and from the U.S. President’s Malaria Initiative (Additional File Section 2).

## Results

### Projected changes in malaria epidemiology

The establishment of *An. stephensi* is predicted to increase the malaria burden across Ethiopia according to pre-existing transmission levels, interventions and vector bionomics. Here we show the increases in prevalence and incidence following introduction and establishment of *An. stephensi* under their current levels of vector-control.

As an example, three locations at very low (∼0.1%), low (∼2.5%) and moderate (∼12%) current *P. falciparum* prevalence are displayed (Figure 2). In all example locations, the introduction of *An. stephensi* leads to increased prevalence, with the most substantial increases in areas with the lowest existing transmission (∼0.1%), though estimates are highly uncertain (Figure 1A). The rate at which prevalence increases depends on the current levels of transmission, with areas at current negligible levels of transmission taking substantially longer than those areas with current moderate-high transmission to reach the new level of endemicity after mosquito establishment. Sub-nationally, there is considerable variation in the projected increases in prevalence, with some administrative groupings experiencing on average minimal (∼0%) increases to absolute prevalence following *An. stephensi* establishment, and others increasing by ∼5.5% (Figure 1B). Overall, the median absolute parasite prevalence increase is ∼2.5% in administrative regions, from ∼1.6% pre-*An. stephensi*. At a national level, we predict a median increase of ∼368,000 (95% CI 103,000 – 664,000) clinical *P. falciparum* malaria cases annually, from a report of ∼740,000 in 2019 [1]. This represents a ∼49.7% (95% CI 13.9 -89.7%) increase if *An. stephensi* only establishes itself in areas previously found suitable and located under 2000m (Figure 1C). Substantially greater increases are seen if *An. stephensi* invades the whole country and can establish malaria transmission at higher altitudes (Additional File Section 3). Temporal changes in clinical incidence over time for the different settings are shown in Figure 2. There is considerable uncertainty in the number of clinical cases following invasion, especially in areas with very low history of malaria where populations have no pre-existing immunity to the disease. Here some model runs projected a substantial increase in malaria cases (greater than in areas with higher malaria prevalence) following a subsequent decline over time that is driven by model assumptions on the acquisition of immunity to clinical disease.

**Figure 1.**
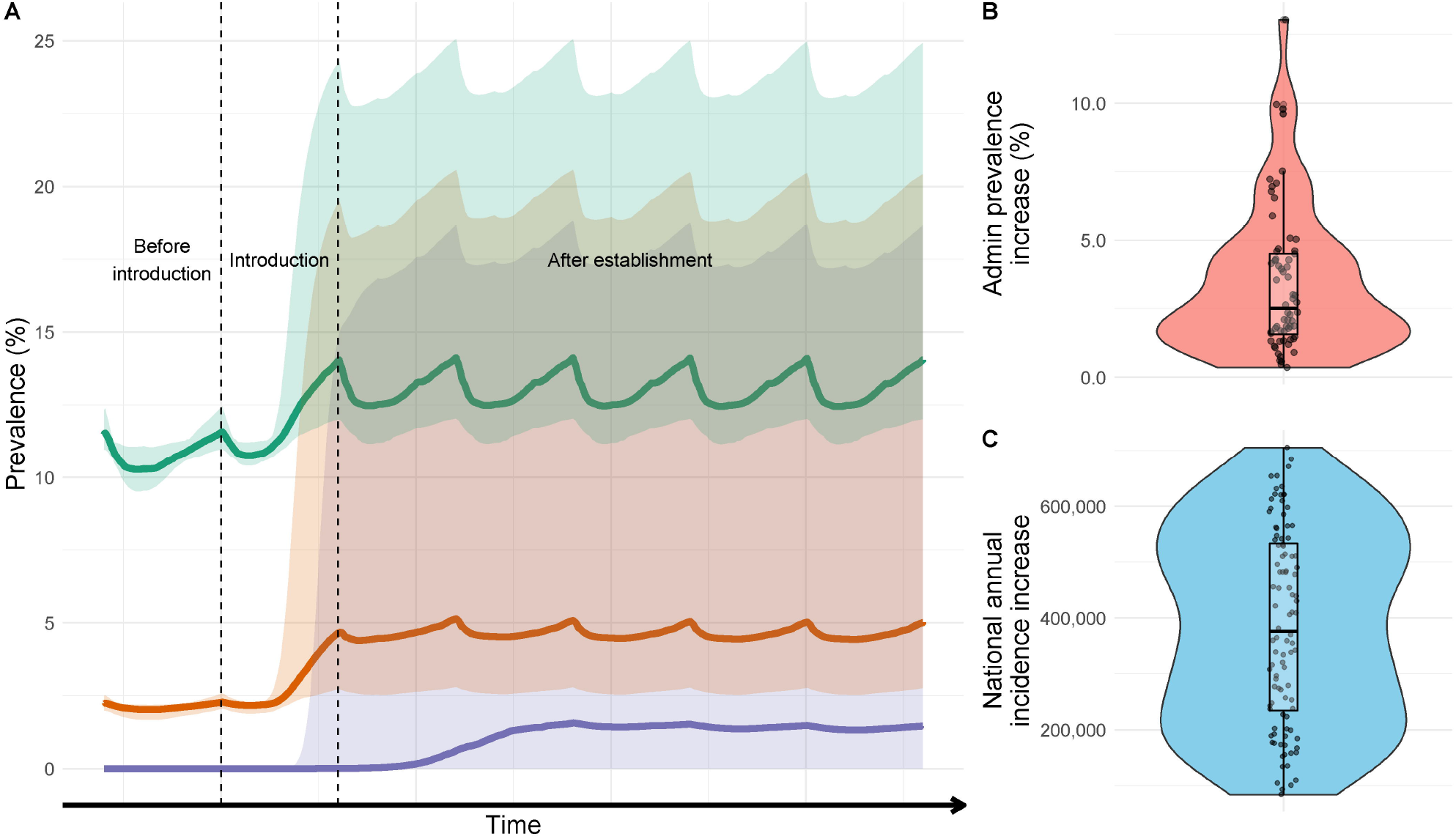
The impact of *An. stephensi* introduction on malaria prevalence and incidence in areas of Ethiopia in areas under 2000m and identified to be suitable. A) Changes in prevalence following introduction and establishment of *An. stephensi* in 3 areas of malaria transmission in Ethiopia. Vertical dashed lines span the period in which *An. stephensi* is introduced and coloured shapes the 95% CI’s. Green represents a transmission scenario of ∼12% prevalence, orange, 2.5%, and purple <0.1%. The time scale on the x-axis is deliberately omitted as the rate of invasion is highly uncertain. Prevalence fluctuating due to pre-existing and ongoing mass distribution of ITNs. B) The median difference of *P. falciparum* prevalence following establishment in the different administrative groupings which have been predicted to be suitable based on Sinka et al., (2020) and under 2000m. C) The annual increase in clinical incidence of malaria caused by *P. falciparum* nationally. Individual dots show uncertainty in predictions given differences in mosquito bionomics (for 100 samples of the Latin hyper cube). Periodicity is caused by ongoing distribution of ITNs and IRS which are assumed to continue at pre-invasion levels. For figure 1B and C there is no x-axis, and points are jittered for visualisation purposes and to aid ease of interpretation rather than to connote a certain value.

**Figure 2.**
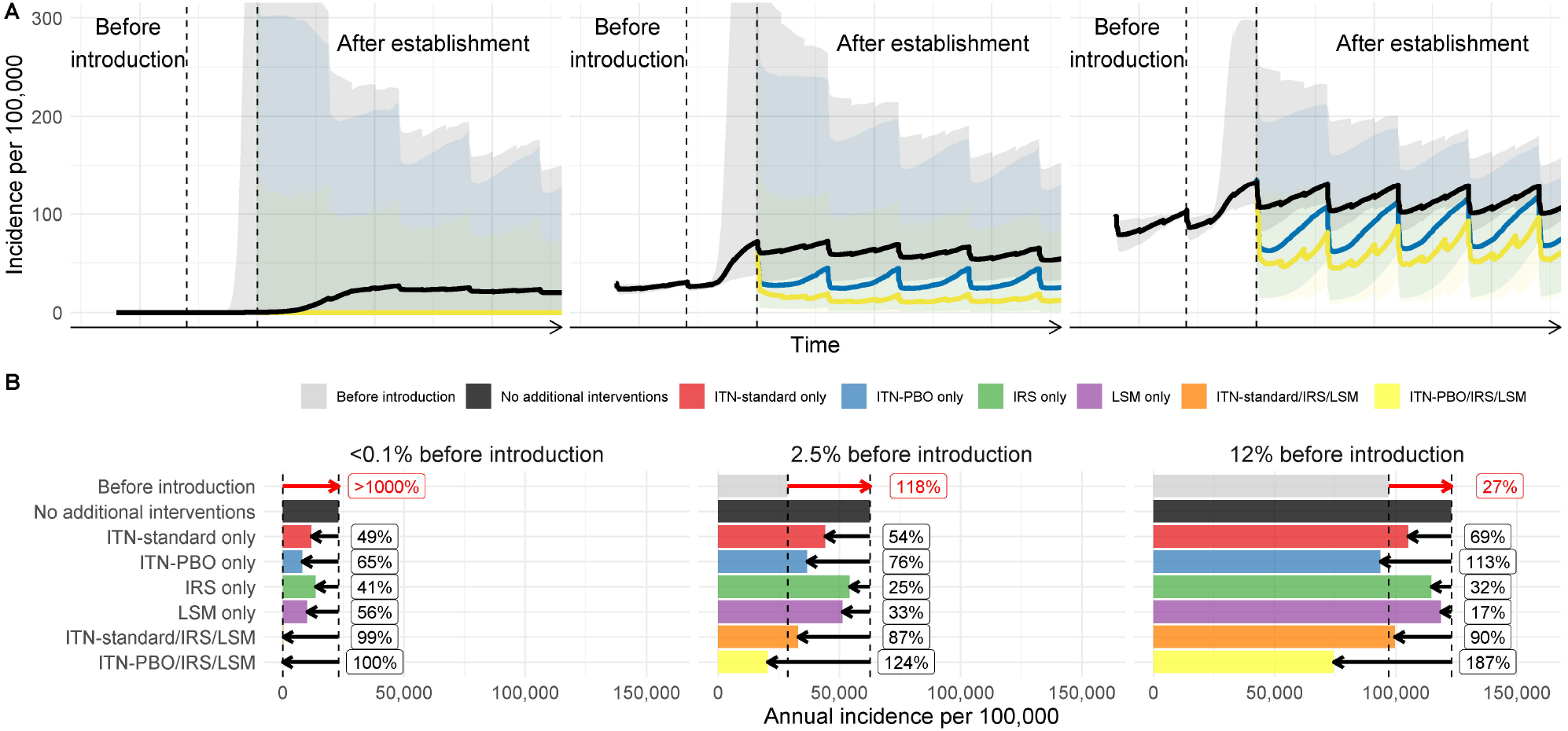
The effect of control measures on mitigating the impact of *An. stephensi* introduction for regions with different pre-existing malaria endemicity. A) The change in clinical incidence over time (per 100,000 people per year) following *An. stephensi* introduction in three locations, with low (∼0.1%), medium (∼2.5%) and high slide prevalence (∼12%). Each location had an EIP of 8 and 40% ITN usage, 0% IRS and 0% LSM pre-*An. stephensi* introduction and establishment. To aid clarity and interpretation of the figure, we only display the temporal trends of the PBO-ITN and PBO-ITN/IRS/LSM package of interventions, though the impacts of all modelled interventions are displayed in Figure 2B. Different coloured lines indicate intervention scenarios be it no additional interventions (black), or scale up of PBO-ITNs (blue), or the use of PBO-ITN/IRS/LSM. Coloured shapes show the 95% CIs for model predictions given uncertainty in parameters. The left vertical dashed line indicates the start to the introduction of *An. stephensi*, which occurs between the dashed lines and is fully established by year 3. As above, the time scale on the x-axis is deliberately omitted due to uncertainty in the timing of events, with clinical incidence fluctuating with 3-year ITN mass campaigns and annual IRS. Additionally, uncertainty continues up to 400 cases per 100,000, but this has been curtailed in order to improve interpretation of median trends. B) The median annual increase in malaria incidence per 100,000 per year comparing before and after scenarios for each of the locations across a range of intervention combinations. Red arrows and numbers refer to the percentage increase in the median incidence following the establishment of *An. stephensi* compared to the 3 years prior to the introduction. Black lines and numbers show the percentage reduction in cases relative to no additional interventions over three years caused by the introduction of the different control interventions.

### Impact of increasing vector control

The impact of additional vector control interventions following *An. stephensi* establishment is dependent on the level of transmission, pre-existing use of interventions and vector bionomics. Mosquito characteristics such as the proportion of bites taken indoors are unknown and can substantially influence the effectiveness of ITNs and IRS (Additional File Section 4). Here we have selected three locations with the same pre-existing intervention usage/coverage and EIP values, but differing levels of pre-existing transmission, in order to illustrate how disease endemicity can influence temporal dynamics (Figure 2). In some circumstances the model predicts that scaling up of standard ITNs, IRS or LSM in combination is sufficient to suppress malaria to pre-*An. stephensi* levels, and sometimes beyond (Figure 2A).

In areas with low levels of transmission (∼0.1%), increases in incidence are substantial, and in order to reduce transmission to pre-*An. stephensi* levels of transmission, a suite of interventions must be considered (Figure 2B). At higher levels of transmission (∼12%), the implementation of PBO-ITNs alone may be suitable to initiate a reduction, however if only pyrethroid-ITNs are available, they should be used in conjunction with IRS and LSM to reduce transmission to pre-*An. stephensi* levels (Figure 2B). The efficacy of LSM in the area is highly unclear, but a 40% reduction in mosquito emergence is predicted to have a larger impact in areas of lower endemicity; conversely, ITNs are predicted to be more effective at higher endemicities. The extent of IRS impact is unclear. Overall, we find that if using standard pyrethroid-only ITNs, a combination of ITN/IRS/LSM is needed to reduce national *P. falciparum* incidence to pre-*An. stephensi* levels. However, given the unknowns in *An. stephensi* responses to these interventions, even this may be insufficient (Table 1, Figure 2B).

**Table 1.** Different combinations of ITN/ITN-PBO/IRS/LSM and the associated annual cases averted and costs, total and per person. ITN/ITN-PBO/IRS values refer to the usage/coverage among the population exposed to *An. stephensi* introduction and LSM the reduction in adult emergence of the established *An. stephensi*. Values in brackets for the cases averted refer to the difference in minimum and maximum 95% CI’s and as such the cases averted median value does not always fall within this, or in numerical order. Total costs and costs per case refer to the median cases averted, and the range in the brackets as the minimum and maximum costs defined in the Additional File.

| ITN | ITN-PBO | IRS | LSM | Cases averted (thousands) | Total costs (million USD) | Cost per case averted (USD) |
| --- | --- | --- | --- | --- | --- | --- |
| 0% |  | 0% | 40% | 68 (9-143) | 9.9 (5.0-14.9) | 146 (556-104) |
| 80% |  | 0% | 0% | 71 (29-31) | 1.8 (1.7-1.9) | 25 (59-61) |
| 0% |  | 80% | 0% | 75 (16-99) | 58.6 (31.7 -84.2) | 781 (1981-851) |
| 80% |  | 80% | 40% | 172 (39-301) | 70.3 (38.4-100.9) | 409 (985-335) |
|  | 80% | 0% | 0% | 139 (60-157) | 3.5 (2.9-3.7) | 25 (48-24) |
|  | 80% | 80% |  | 207 (70-33) | 64.6 (37.2-90.8) | 312 (531-2752) |
|  | 80% |  | 80% | 210 (49-361) | 16.0 (10.6-21.5) | 76 (216-60) |
|  | 80% | 80% | 40% | 302 (79-879) | 72.0 (39.9-102.5) | 238 (512-117) |

The mass distribution of PBO-pyrethroid nets may offer a more cost-effective tool given the high levels of pyrethroid resistance and relatively low levels of endophily in Ethiopian *An. stephensi* populations (Figure 2 and Table 1). PBO-pyrethroid nets are predicted to have the most substantial impact on transmission of any single intervention examined, and almost achieves the reduction provided by a combination of standard ITNs, IRS and LSM (139,000 (60,000-157,000) vs 194,000 (44,000-410,000) cases averted). Furthermore, PBO-pyrethroid nets only account for a fraction of the cost of this more comprehensive intervention packet, $3.5 (2.9-3.7) vs $70.3 million USD (38.4-100.9).

Layering multiple vector control interventions are predicted to have the largest impact. However, because of the number of sites with minimal malaria transmission before *An. stephensi* invasion, this may be insufficient to push the national malaria burden beneath current levels unless PBO-pyrethroid nets, higher usage/coverage (ITNs, IRS) or improved effectiveness (LSM) can be achieved (Table 1). Increases in IRS coverage to 80% alone offers substantial improvements due to the relatively limited use of IRS currently nationwide (Additional File Section 2). However, due to low rates of endophily and crepuscular biting, the impact is substantially less than would be expected against African *Anopheles* species. The cost of implementing interventions is expected to be substantial, with the most comprehensive set of interventions (80% use of PBO-pyrethroid ITN and IRS, 40% reduction in adult emergence) estimated to cost an additional $72.0 million USD ($39.9 -$102.5 million) annually or $238 USD ($117-$505) per case averted (over existing malaria prevention budgets).

## Discussion

The invasion and establishment of *Anopheles stephensi* represents an imminent and substantial threat to the Horn of Africa and wider region which could jeopardise progress achieved in malaria control. The modeled increase in malaria is highly uncertain, but without additional interventions, the impact could be considerable, with an estimated ∼50% increase from the reported ∼740,000 *P. falciparum* malaria cases in Ethiopia, 2019, to an estimated 1,130,000 (95% CI 843,000-1,404,000) after establishment and disease equilibrium has been reached. This assumes that malaria cases only increase in areas previously predicted as suitable for *An. stephensi* establishment located under 2000m. Numbers could be substantially higher if invasion is more widespread.

Sub-nationally, significant heterogeneity in public health impact is expected. Analysis suggests that low altitude urban areas at pre-existing low levels of malaria transmission may experience the largest increases in disease burden. In these areas, the absence of existing vector control and low human population immunity indicates the possibility for substantial increases in transmission. These areas are also the most uncertain, with the model predicting the increase that clinical cases could be negligible to more than that observed in areas where malaria has historically been widespread. Our finding that areas with low pre-existing levels of transmission (much of Ethiopia) take substantially longer to see increases in malaria after *An. stephensi* introduction is particularly worrying. In these locations, in the absence of widespread and routine surveillance for *An. stephensi*, the first signal detected could be an increase in malaria, which would occur a considerable amount of time after mosquito establishment. While this would not be of significant concern if the vector was easily removed, or only led to a relatively minor and easily combatable increase in malaria, our model findings and what has been observed in Djibouti [11, 12] suggest that this is not the case. Once *An. stephensi* becomes established, it could lead to significant increases in malaria transmission that are not easily reversed. This unnoticed proliferation and subsequent increase in transmission was previously seen in *Anopheles arabiensis* in North-Eastern Brazil, where its “silent spread” led to a large malaria epidemic [31, 63]. Nevertheless, this work shows that the absence of an increase in reported malaria cases following the identification of the presence of *An. stephensi* should not be overly interpreted.

To combat the possible increase of malaria transmission following *An. stephensi* establishment, the deployment of a wide array of vector control interventions should be considered. High levels of pyrethroid resistance observed in *An. stephensi* captured in Ethiopia [17, 18] suggest that pyrethroid-only ITNs (already in use across the country) will have a limited efficacy for control of malaria transmitted by *An. stephensi*. Adoption of PBO-ITNs is predicted to be both highly impactful and cost-effective. The impact of different control interventions is unclear given the receptiveness of *An. stephensi* in this new environment is unquantified. For example, if the mosquito feeds at night and rests inside structures sprayed with IRS then the widespread use of this intervention alone could be sufficient to mitigate the public health impact (Additional File Section 4). However, if the mosquito was less amenable to indoor vector control, then layering of ITNs and IRS may be insufficient. Within its endemic range, *An. stephensi* shows a propensity to both crepuscular biting and resting outside of houses compared to African *Anopheles* species, suggesting a reduced efficacy if it behaves as it does in its endemic range [31] (Additional File Section 2). Compounding this, high-density urban locations, where large scale vector-control campaigns have been historically absent, will present a challenge for establishing ITN access and use, as well as achieving sufficient IRS coverage. While *An. stephensi* is primarily known as an urban vector of malaria, it is found in both urban and rural settings across its endemic range [31], and in Ethiopia [18], and so there is the potential for its impact on malaria transmission to be found across the country. Differences in environment, housing, culture, human and vector behaviour in urban and rural settings are likely to result in very different public health outcomes, even before considering the logistics of intervention deployment. Though these factors are likely to have a substantial effect, it is presently unknown how these combined factors will translate into malaria transmission and control. In the absence of quantified and validated input data we have not stratified models by urbanicity at this present time. Nevertheless, additional data on *An. stephensi* bionomics and the impact of vector control interventions, stratified across urban/rural areas and housing types, should therefore be collected as a matter of urgency to enable model estimates to be refined and contribute to the decision-making process, (Additional File Section 4). These compound uncertainties combined with the crude method of economic evaluation adopted here make cost projections highly variable. Though these are substantial ($74.6 million USD ($42.6 - $105.4)), the economic burden of an additional ∼368,000 (95% CI 103,000 – 664,000) malaria cases should not be underestimated.

This work was intended to provide initial estimates of the possible scale and impact of *An. stephensi* invasion rather than detailed predictions of what will happen. As such, there are many limitations to this approach (further detailed in the Additional File Section 4)and results should not be overly interpreted. The public health impact is likely to be underestimated as we only consider *P. falciparum* malaria, despite *An. stephensi* having been shown to be capable of efficiently transmitting *P. vivax* in Ethiopia [59]. Due to the presence of additional *Anopheles* species in Djibouti, it is unlikely the increases we have seen are purely due to *An. stephensi*, which we have assumed, and while highly correlated [11], and increasingly implicated [12], the causative role in *An. stephensi* in the increases seen in Djibouti has not been established. However, the trends observed along with evidence of underreporting in Djibouti are cause for significant concern [12]. Malaria burden is related to the extent to which *An. stephensi* may invade a region (i.e., the number of mosquitoes per person and not just its presence), and the seasonality of the vector. Malaria transmission is often highly seasonal, and an important factor when evaluating local epidemiology and vector-control. However, the case of *An. stephensi* seasonality is a complicated and currently unquantified unknown. Across its endemic range, *An. stephensi* shows a variety of patterns from unimodal to bimodal to peaking in the dry season to the wet [64], and within Ethiopia there is a dearth of this data. As we are not modelling seasonally timed interventions, and are concerned with long term rather than intra-annual dynamics, the omission of seasonality in our model does not change model predictions or recommendations, though the quantification of *An. stephensi* seasonality in the Horn of Africa should be a matter of urgent study.

## Conclusions

While we have made use of available published and unpublished sources on *An. stephensi* bionomics, there is either insufficient data or substantial intra-species variability to simply ascribe a set of characteristics to how *An. stephensi* will interact with humans and control interventions in Ethiopia. In order to improve predictions, we have identified priority aspects of *An. stephensi* bionomics (such as anthropophagy, endophily, indoor biting), local transmission (existing distribution of malaria cases between rural and urban areas) and intervention parameters (efficacy of ITNs, IRS and LSM give a certain effort) that should be explored as a priority in order to inform additional mathematical modelling of the impact of *An. stephensi* on malaria transmission in Africa (see Additional Section 4).

Though there is substantial uncertainty in what will happen if *An. stephensi* becomes established in Ethiopia and other locations across Africa, the estimates provided here, and the situation seen in Djibouti, should be a stark warning against complacency and highlight the need to rapidly improve surveillance and evaluate effective control interventions in response to this developing threat.

## Data Availability

All data and code required to run the analyses are found at https://github.com/arranhamlet/stephensi_ETH_publication

## Abbreviations

ITN: Insecticide treated nets
IRS: Indoor residual spraying
PBO: piperonyl butoxide
EIP: Extrinsic Incubation Period
LHS: Latin Hypercube Sampling
MAP: Malaria Atlas Project
WMR: World Malaria Report
LSM: Larval Source Management
PMI: Presidents Malaria Initiative
WHO: World Health Organization

## Declarations

### Ethics approval and consent to participate

Not applicable, as all data used was publicly available and modelled rather than individual data.

### Consent for publication

Not applicable

### Availability of data and materials

All data and code required to run the analysis is provided at https://github.com/arranhamlet/stephensi_ETH_publication [65]

## Acknowledgements

NA

## Authors contributions

Conceptualization: A.H, T.C, A.S, S.R.I, J.S.A

Methodology; A.H, T.C

Software: A.H.

Supervision: T.C, A.S, S.R.I, J.S.A

Project administration: T.C, A.S, S.R.I, J.S.A

Data curation: A.H, J.E.T, A.S, S.R.I

Writing: A.H, D.D, J.E.T, F.G.T, T.B, M.S, A.S, S.R.I, J.S.A, T.C

Reviewing: A.H, D.D, J.E.T, F.G.T, T.B, M.S, A.S, S.R.I, J.S.A, T.C

Visualisation: A.H

All authors read and approved the final manuscript

## Additional file description

Further details of the mechanistic model and underlying data, further analyses and additional limitations and highlighting useful data for collection to further mathematical modelling. Figure S1 - Mechanistic model schematic. Figure S2 - Parameters included for each administrative grouping. Figure S3 – Relationship of EIP and temperature. Table S1 – Mosquito bionomics. Table S2 – Grouping of administrative units and values. Table S3 – Cost per intervention per person per year. Figure S4 – Vector bionomics and fitted incidence to data. Figure S5 – Sensitivity analysis on impact across assumptions of population exposed. Table S4 – Populations and increases in clinical incidence across assumptions of population exposed. Figure S6 – Relationship of prevalence and EIP. Table S5 – Key parameters to inform modelling and impacts. Figure S7 – Univariate analysis of entomological parameters and incidence. Table S6 – Vector bionomics from literature search.

