## Supplementary information for "The potential impact of *Anopheles stephensi* establishment on the transmission of *Plasmodium falciparum* in Ethiopia and prospective control measures"

**Additional information**

### **1. Mechanistic model**

Here we use a deterministic version of a well-established and highly utilised compartmental model of *Plasmodium falciparum* malaria transmission^19, 20, 22, 23, 25^, which models transmission within humans and vectors at various stages of infection. We account for heterogeneity in transmission as well as age-dependent biting rates and the acquisition of natural immunity. The model has previously been described fully in the aforementioned publications, but we summarise it briefly below (Additional File Figure 1).

When Susceptible (S) individuals become infected, they progress to either an asymptomatic (A) state or clinical disease, at a rate dependent on the force of infection, Λ, and the probability of acquiring clinical disease, φ, which is itself dependent on natural immunity. Those progressing to clinical disease either enter the treated (T) or clinical disease (D) compartment dependent on the probability of treatment (f_T_). Treated individuals progress through to a period of protection through prophylaxis (P), at rate r_T_, and return to the susceptible compartment at rate r_P_.

Individuals in clinical disease (D) remain symptomatic for the duration of the disease course, r_D_, then move to an asymptomatic state (A), which is detectable through microscopy, before the infection becomes submicroscopic (so undetectable through microscopy, U) at rate r_A_. Asymptomatic individuals (in either the A or U compartment) can develop clinical disease, but if they do not, they clear the infection and return to the symptomatic compartment at rate r_U_. Adult mosquito populations are modelled through a susceptible (S­_m_) and progress to the exposed (E_m_) state at rate Λ_m_, and onto the infectious (I_m_) after the extrinsic incubation period (EIP) has been completed. Mosquitoes are exposed to human infection through feeding through the treated (T), clinical disease (D), asymptomatic (A) and submicroscopic (U) infection states.

Vector control interventions are assumed to reduce malaria by primarily killing adult mosquitoes but also through dissuading them from biting. The use of insecticide treated nets (ITNs) is incorporated in the model structure using the methods outlined in Le Menache et al., 2007^[26]^ and Griffin et al. 2010^[21]^. Entomological data has shown that ITNs are less effective against pyrethroid resistant mosquitoes (as described by the percentage of mosquitoes surviving a discriminating dose bioassay). The efficacy of ITNs in Ethiopia is calculated using local discriminating dose bioassay data and methods outlined by Churcher *et al* (2016)^[27]^. The efficacy of indoor residual spraying (IRS) was parameterised using Sherrard-Smith et al 2018 whilst larval source management was assumed to reduce mosquito emergency by a defined value constantly throughout the year. ITNs reduce human biting by repelling, and killing mosquitoes before biting, while IRS can either repel before biting and kill after biting (when the vector rests on an inside wall). Furthermore, both ITN and IRS efficacy decays over its lifespan due to a loss of insecticide (Churcher et al. 2016)^[27]^. The supplementary material of Walker (2016) further elaborates on these mechanisms^[28]^.

Models were parameterised for each site individually taking into account the history of control interventions (see main text for a description and Figure A2 for values). The human-to-mosquito ratio was varied to match the observed malaria prevalence in the defined cohort. To simulate the invasion of *Anopheles stephensi*, the vector density is increased in a sigmoidal fashion over 3 years to approximately match the patterns seen in malaria incidence in Djibouti.

The package required to run the model and a version of the model code is found at <https://github.com/mrc-ide/deterministic-malaria-model>.

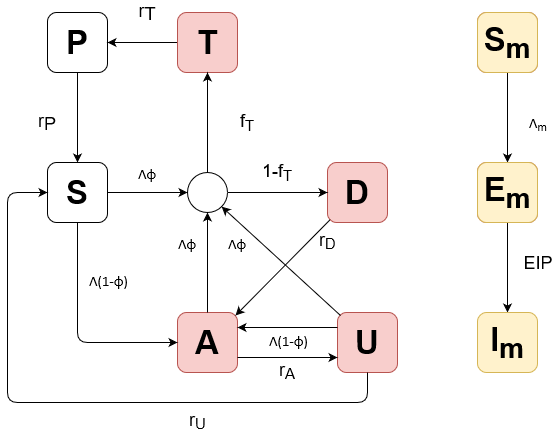

**Additional File Figure 1. Model diagram showing the progression between human and vector states.** Humans are either S = susceptible, A = asymptomatic, T = treated, D = have clinical disease, or U = submicroscopic infection, whereas mosquitoes are either S_m_ = Susceptible, E_m_ = infected but not infectious, I_m_ = infectious. The arrows shown transitions between compartments and the circle represents treatment. Red compartments indicate states that can expose susceptible mosquitoes to infection and yellow the mosquito compartments. The life-cycle of the pre-adult life-stages of the mosquito are omitted for simplicity though see ^4^ for full details.

### **2. Data**

#### **Human population sizes and pre-existing malaria prevalence**

Administrative unit human population sizes were obtained by using WorldPop 2020 population raster, which provides population estimates at a 1/120 degree resolution^[29]^. This was then standardised to the 2020 Ethiopia country level population estimate from the UN World Population prospects^[30]^. This raster was then applied to the administrative boundaries in order to estimate populations in each unit.

To estimate the population below a certain altitude or within areas found suitable by previous research^[31]^, we applied theses limits to the above standardised population raster using a suitability raster provided by Sinka et al., (2020)^[31]^ and altitude from WorldClim^[32]^. Malaria prevalence, *P. falciparum*, in 2-10-year olds is provided by the Malaria Atlas Project (MAP) and aggregated to the administrative level by taking the population weighted mean.

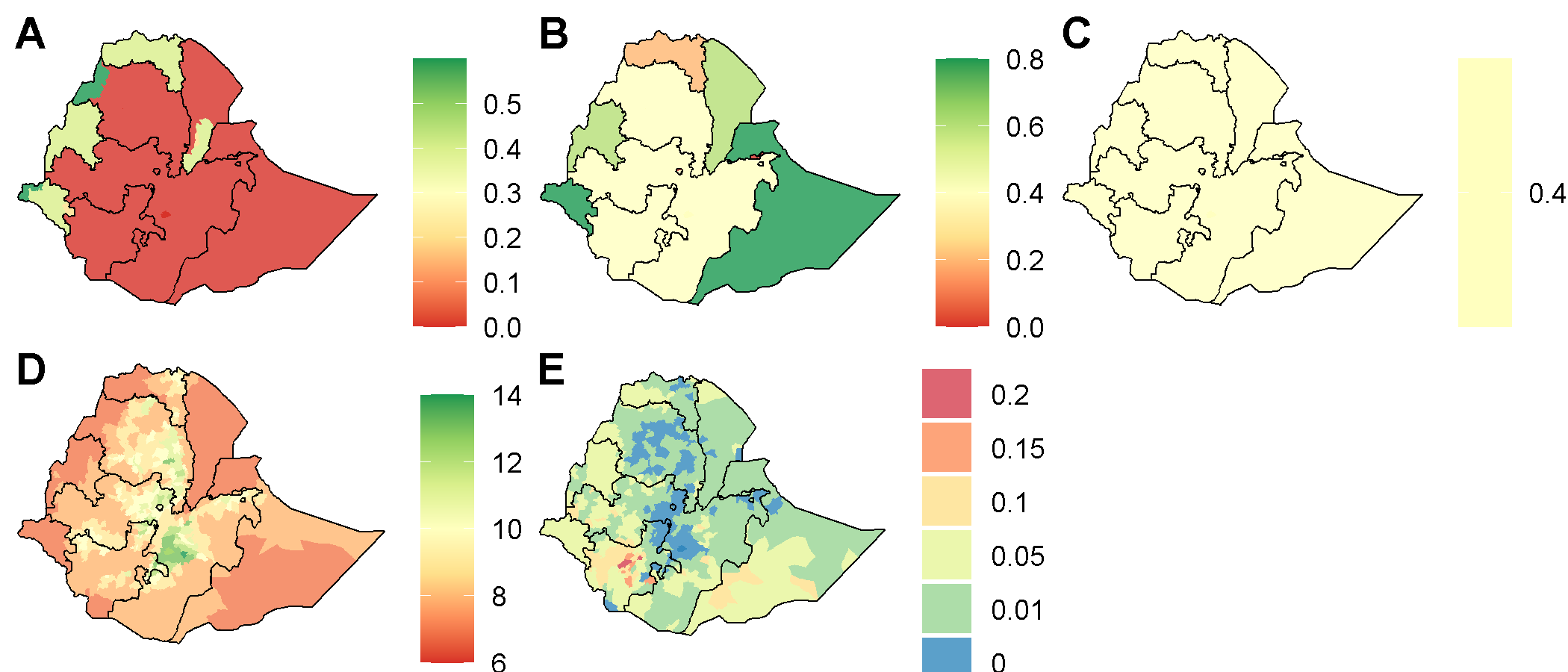

**Additional File Figure 2. Parameters included for each administrative grouping.** A) IRS coverage (%), B) ITN coverage (%), C) treatment coverage (%), D) extrinsic incubation period (days), E) Prevalence in 2-10-year-olds.

#### **Temperature and EIP estimates**

Temperature data was accessed by WorldClim^[32]^ which provided monthly temperature data at the maximum, mean and minimum for 2010-2018.

Highland regions of Ethiopia are predicted to be too cold to sustain malaria transmission throughout the year. As a result, here we have used the hottest months mean maximum temperature in the year, as it represents the “extreme” scenario – with the fastest EIP. While this may overestimate the value of EIP overall, the majority of malaria transmission occurs in relatively short “transmission seasons”, and so it only takes a few months of heightened suitability in an otherwise less suitable temperature clime to have a substantial effect. This was then averaged over the period to produce a single raster file of temperature at a resolution of 1/120 degrees.

This temperature was then converted into EIP estimates based on a previous quantification by Stopard et al., (2021) which provided a description of the temperature dependent relationship of *P. falciparum* in *An. Stephensi*^[33]^ (Additional File Figure **3**). These estimates were derived in laboratory populations of mosquitoes and parasites though is assumed to hold for wild *An. stephensi* mosquitoes infected with *P. falciparum* in Ethiopia^[34-37]^. For simplicity it is assumed that all mosquitoes have the same EIP. As this temperature relationship was modelled on a limited set of temperatures (21-34 °C) it was necessary to extrapolate this further to capture the temperature range of Ethiopia. This EIP estimate was converted to the administrative level by taking the mean EIP in an area (Additional File Figure **2D**). All other mosquito bionomics and factors influencing transmission biology are assumed to be independent of temperature or altitude.

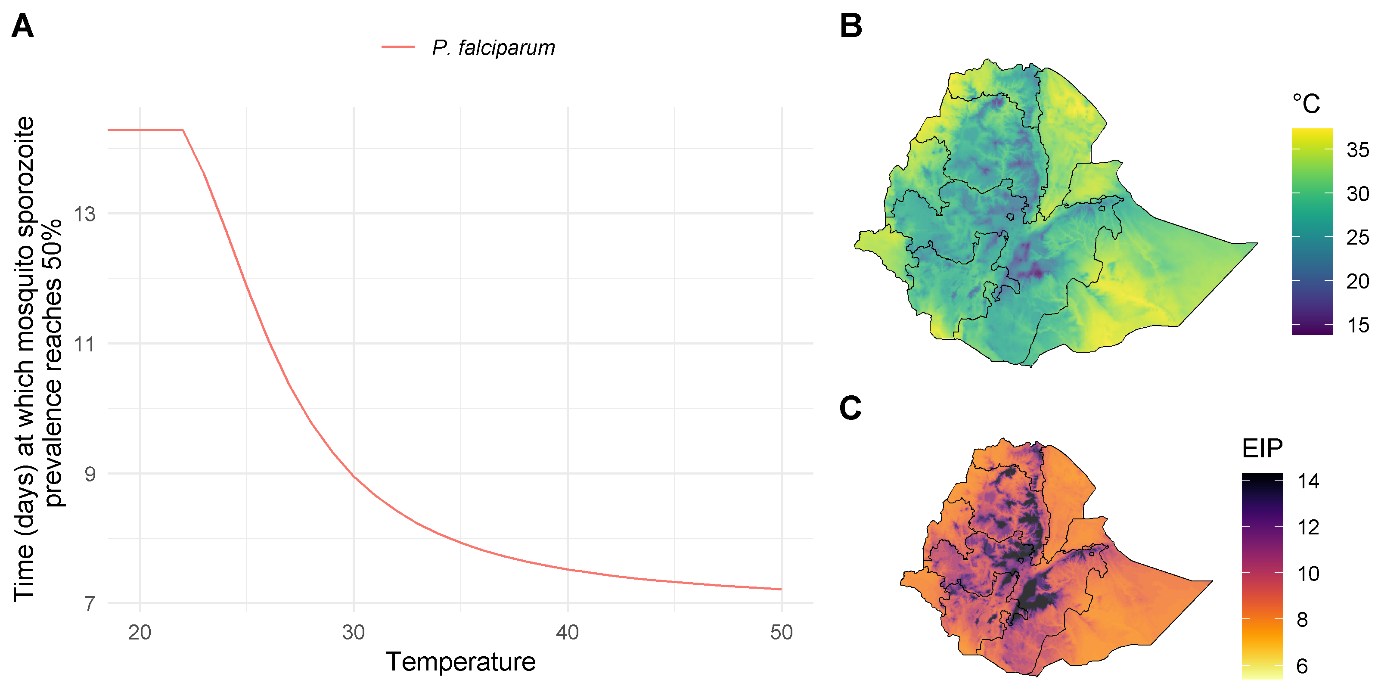

**Additional File Figure 3. How parasite extrinsic incubation period is assumed to vary across Ethiopia.** A) Relationship between the extrinsic incubation period and temperature for the development of *Plasmodium falciparum* in *Anopheles stephensi* in laboratory studies. B) The maximum monthly annual temperature across Ethiopia. C) Predicted EIP (days) derived from the maximum temperature for *P. falciparum.*

#### **Mosquito bionomics**

The results of a literature search for the key mosquito bionomics parameters are summarised in Additional File Table 1. Data re relatively scant and those studies that were identified show *An. stephensi* estimates vary substantially in its native range. In addition, very few studies were identified within the context of Djibouti or Ethiopia. To account for this uncertainty a wide range of parameter distributions were used which are presented in (Additional File Figure **4**A).

#### **Shapefiles and grouping administrative units**

Shapefiles were provided through the humanitarian data exchange (<https://data.humdata.org/dataset/ethiopia-cod-ab>) at the country, 1^st^, 2^nd^ and 3^rd^ administrative unit level. Administrative units at the 3^rd^ level were grouped based on their pre-existing transmission, interventions and EIP. Here we round prevalence to the nearest 5%, with an additional value of 1% as to capture areas with low but not negligible levels of transmission which account for much of Ethiopia. Interventions and treatment coverages were rounded to the nearest 20% and EIP to the nearest integer. This rounding allows us to substantially reduce the number of runs required, and by using approximate rather than “exact” values from the data we aim to not overstate the accuracy of our findings. By then grouping administrative locations by their combination of parameters, we can reduce the number of simulations required for each model run from 690 (each individual adm3 location) to 64. Full details of the diversity of groups are shown in Additional File Table 3.

**Additional File Table 1. Mosquito parameters and values used in Latin hypercube sampling.**

| Parameter | Values |
| --- | --- |
| Daily mortality | 0.093 – 0.154 |
| Proportion of blood meals taken on humans,  anthropophagy | 0.1 – 0.4 |
| Proportion of mosquitoes resting inside, endophily | 0.375 – 0.625 |
| Proportion of mosquito bites taken when people indoors (in absence of interventions) | 0.358 – 0.597 |
| Proportion of mosquito bites taken when people in bed (in absence of control interventions) | 0.391 – 0.652 |

**Additional File Table 2. Vector bionomics taken from literature search.**

| Source | country | urban/rural | parameter | value |
| --- | --- | --- | --- | --- |
| 38 | Iran | rural | proportion of anthropophagy | 0.278 |
| 38 | Iran | rural | proportion of anthropophagy | 0.183 |
| 38 | Iran | rural | proportion of anthropophagy | 0.35 |
| 38 | Iran | rural | proportion of anthropophagy | 0.157 |
| 38 | Iran | urban | proportion of anthropophagy | 0.435 |
| 39 | Iran | rural | proportion of anthropophagy | 0.118 |
| 40 | Iran | rural | proportion of anthropophagy | 0.1 |
| 41 | Iran | rural | proportion of anthropophagy | 0.27 |
| 41 | Iran | rural | proportion of vector endophily | 0.6125 |
| 42 | Iran | rural | endophagy in bed | 0.457333333 |
| 5 | India | urban | proportion of vector endophily | 0.065408805 |
| 5 | India | urban | proportion of anthropophagy | 0.009124088 |
| 43 | Iran | rural | proportion of anthropophagy | 0.1983 |
| 44 | India | urban | proportion of anthropophagy | 1 |
| 45 | India | unknown | duration of host-seeking behaviour | 3 |
| 46 | Iran | rural - mountainous | proportion of vector endophily | 0.518 |
| 46 | Iran | rural - plains | proportion of vector endophily | 0.538 |
| 47 | Pakistan | rural | proportion of vector endophily | 0.80296631 |
| 47 | Pakistan | rural | proportion of vector endophily | 0.603058104 |
| 47 | Pakistan | rural | proportion of vector endophily | 0.881918819 |
| 48 | Laboratory | NA | daily mortality of adult mosquitoes | 0.047619048 |
| 49 | Pakistan | rural | proportion of anthropophagy | 0.024 |
| 49 | Pakistan | rural | proportion of anthropophagy | 0.016 |
| 49 | Pakistan | rural | daily mortality of adult mosquitoes | 0.304878049 |
| 49 | Pakistan | rural | daily mortality of adult mosquitoes | 0.266666667 |
| 50 | Kolkata | urban | proportion of vector endophily | 0.634408602 |
| 50 | Kolkata | urban | endophagy indoors | 0.676 |
| 51 | Laboratory | NA | daily mortality of adult mosquitoes | 0.12345679 |
| 52 | India | urban - riverine | proportion of anthropophagy | 0.0045 |
| 52 | India | urban - non-riverine | proportion of anthropophagy | 0.014 |
| 52 | India | urban | proportion of anthropophagy | 0.0103 |
| 52 | Iran | urban | proportion of anthropophagy | 0.086 |
| 52 | Iran | urban | proportion of anthropophagy | 0.049 |
| 53 | India | urban | proportion of vector endophily | 0.214728879 |
| 54 | Iran | rural | proportion of vector endophily | 0.38976378 |
| 54 | Iran | rural | proportion of anthropophagy | 0.2375 |
| 55 | Ethiopia | rural | proportion of anthropophagy | 0.234 |

**Additional File Table 3. Grouping of administrative units and the number of admin units per category (total 650), by *P. falciparum* prevalence (%), IRS/ITN coverage (%) and EIP (days).**

| ID | No. admin units | P. falciparum prevalence (2-10 years) | IRS | ITN | EIP |
| --- | --- | --- | --- | --- | --- |
| 1 | 1 | 0 | 0% | 40% | 14 |
| 2 | 11 | 0 | 0% | 40% | 13 |
| 3 | 2 | 0 | 0% | 0% | 12 |
| 4 | 22 | 0 | 0% | 40% | 12 |
| 5 | 4 | 0 | 0% | 0% | 11 |
| 6 | 28 | 0 | 0% | 40% | 11 |
| 7 | 1 | 0 | 40% | 20% | 10 |
| 8 | 26 | 0 | 0% | 40% | 10 |
| 9 | 3 | 0 | 40% | 20% | 9 |
| 10 | 31 | 0 | 0% | 40% | 9 |
| 11 | 1 | 0 | 40% | 20% | 8 |
| 12 | 15 | 0 | 0% | 40% | 8 |
| 13 | 1 | 0 | 0% | 80% | 8 |
| 14 | 1 | 0 | 0% | 40% | 7 |
| 15 | 3 | 0 | 0% | 60% | 7 |
| 16 | 2 | 0.01 | 0% | 40% | 13 |
| 17 | 3 | 0.01 | 0% | 40% | 12 |
| 18 | 11 | 0.01 | 0% | 40% | 11 |
| 19 | 3 | 0.01 | 40% | 20% | 10 |
| 20 | 50 | 0.01 | 0% | 40% | 10 |
| 21 | 5 | 0.01 | 40% | 20% | 9 |
| 22 | 68 | 0.01 | 0% | 40% | 9 |
| 23 | 2 | 0.01 | 0% | 80% | 9 |
| 24 | 2 | 0.01 | 0% | 0% | 8 |
| 25 | 11 | 0.01 | 40% | 20% | 8 |
| 26 | 117 | 0.01 | 0% | 40% | 8 |
| 27 | 3 | 0.01 | 40% | 60% | 8 |
| 28 | 13 | 0.01 | 0% | 80% | 8 |
| 29 | 3 | 0.01 | 40% | 20% | 7 |
| 30 | 4 | 0.01 | 0% | 40% | 7 |
| 31 | 2 | 0.01 | 60% | 40% | 7 |
| 32 | 19 | 0.01 | 0% | 60% | 7 |
| 33 | 1 | 0.01 | 20% | 60% | 7 |
| 34 | 8 | 0.01 | 40% | 60% | 7 |
| 35 | 12 | 0.01 | 0% | 80% | 7 |
| 36 | 6 | 0.05 | 0% | 40% | 11 |
| 37 | 2 | 0.05 | 0% | 40% | 10 |
| 38 | 1 | 0.05 | 40% | 20% | 9 |
| 39 | 28 | 0.05 | 0% | 40% | 9 |
| 40 | 1 | 0.05 | 40% | 20% | 8 |
| 41 | 59 | 0.05 | 0% | 40% | 8 |
| 42 | 2 | 0.05 | 0% | 60% | 8 |
| 43 | 6 | 0.05 | 40% | 60% | 8 |
| 44 | 4 | 0.05 | 0% | 80% | 8 |
| 45 | 2 | 0.05 | 20% | 80% | 8 |
| 46 | 5 | 0.05 | 40% | 20% | 7 |
| 47 | 3 | 0.05 | 0% | 40% | 7 |
| 48 | 1 | 0.05 | 60% | 40% | 7 |
| 49 | 2 | 0.05 | 0% | 60% | 7 |
| 50 | 8 | 0.05 | 40% | 60% | 7 |
| 51 | 16 | 0.05 | 0% | 80% | 7 |
| 52 | 7 | 0.05 | 40% | 80% | 7 |
| 53 | 4 | 0.05 | 60% | 80% | 7 |
| 54 | 1 | 0.1 | 0% | 40% | 11 |
| 55 | 2 | 0.1 | 0% | 40% | 10 |
| 56 | 6 | 0.1 | 0% | 40% | 9 |
| 57 | 21 | 0.1 | 0% | 40% | 8 |
| 58 | 1 | 0.1 | 0% | 80% | 8 |
| 59 | 1 | 0.1 | 0% | 40% | 7 |
| 60 | 4 | 0.1 | 0% | 80% | 7 |
| 61 | 1 | 0.15 | 0% | 40% | 10 |
| 62 | 1 | 0.15 | 0% | 40% | 9 |
| 63 | 4 | 0.15 | 0% | 40% | 8 |
| 64 | 2 | 0.2 | 0% | 40% | 8 |

#### **Cost of interventions per person**

Approximate estimates of the cost of intervention (purchasing, delivering, and applying) were provided from literature and from the PMI through personal communication. We have assumed 1.8 people per ITN, which were assumed to be standard pyrethroid nets.

**Additional File Table 4. Approximate estimated costs per person per year for the different vector control interventions considered.**

| Intervention | Costing estimate per year per person | | |
| --- | --- | --- | --- |
|  | Low | Medium | High |
| ITN-PBO-pyrethroid | $0.56 | $0.61 | $0.66 |
| ITN-pyrethroid | $0.43 | $0.45 | $0.49 |
| IRS | $3.35 | $6.19 | $8.9 |
| Larvicide | $1.00 | $2.00 | $3.00 |

### **3. Additional analyses**

#### **Model fit to Djibouti data**

A hundred models varying mosquito bionomics were fit to the MAP estimates of annual clinical incidence in Djibouti (Additional File Figure 4B). Anthropophagy refers to the proportion of blood meals taken on humans, endophagy bed refers to the proportion of blood meals taken in bed, endophagy indoors the proportion of blood meals taken indoors, endophily the proportion of time spent around human houses and mortality, the daily probability of an adult mosquito dying. Overall, there was a very high rate of agreement between the model estimates and the data, with point estimates approximately equally and well within 95% CI’s.

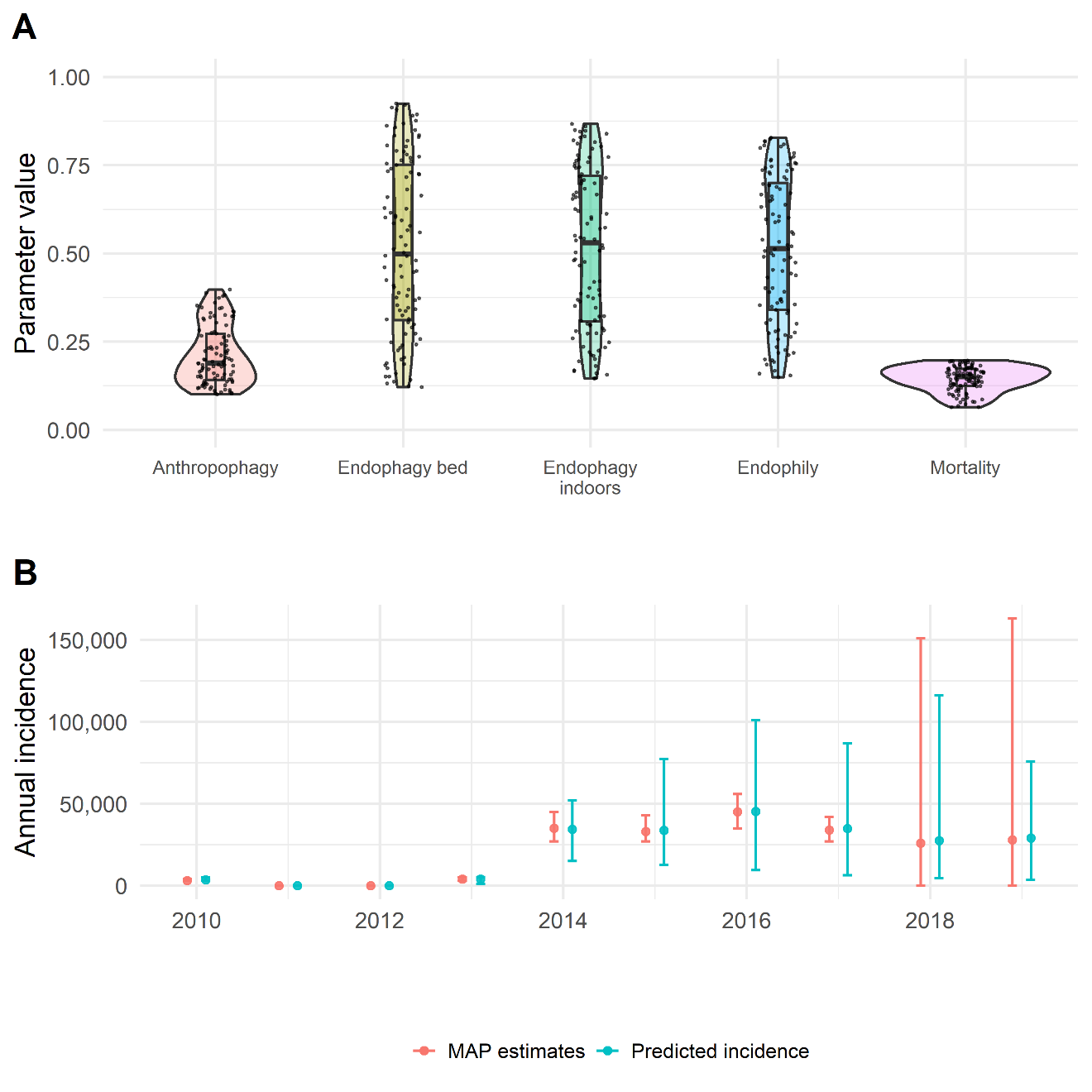

**Additional File Figure 4. Vector bionomics parameter distributions and fitted model incidence compared to the MAP estimates calibrated against**. A) The distribution of the parameters used from the 100 best LHC models. Violin plots show density of data, the boxplot the quantiles and outliers and the points the individual values. B) The model predicted incidence against the MAP incidence the model was calibrated against, error bars show the 95% CI’s. The vector densities required to produce these estimates were a median of 6.8 (95% CI 3.2 - 35.1).

#### **Impact of *Anopheles stephensi* under different assumptions of populations exposed.**

Here we have varied where *An. stephensi* is predicted to increase based on pre-existing estimates of regional suitability for *An. stephensi* as estimated using the geostatistical models of ^11^. This particularly highlights urban areas, though many people reside in these regions meaning the epidemiological impact could be substantial (Additional File Figure **5**). Another method of generating alternative metrics of the population suitable for ongoing *falciparum* malaria transmission is to only include communities under 2000m, as estimate of an altitude above which malaria transmission is less suitable. This assumption means that a much wider geographical region is suitable but excludes some large cities so the population at risk is substantially lower (**Additional File Table 5**).

If *An. stephensi* is confined to areas previously deemed as suitable, then we project an additional 531,000 (134,000 - 979,000) malaria cases a year, and 1,884,000 (542,000 - 3,348,000) if we consider anywhere under 2000m to be suitable. In the most extreme scenario, assuming the entire country is suitable regardless of altitude or prior prediction of suitable, we estimate 3,163,000 (818,000 - 5,740,000) additional malaria cases per year (Additional File Figure 5 and **Additional File Table 5**). In all scenarios, these estimates of additional annual malaria cases are substantially larger than the baseline assumption of only areas under 2000m and previously indicated as suitable, of 368,000 (103,000 - 664,000).

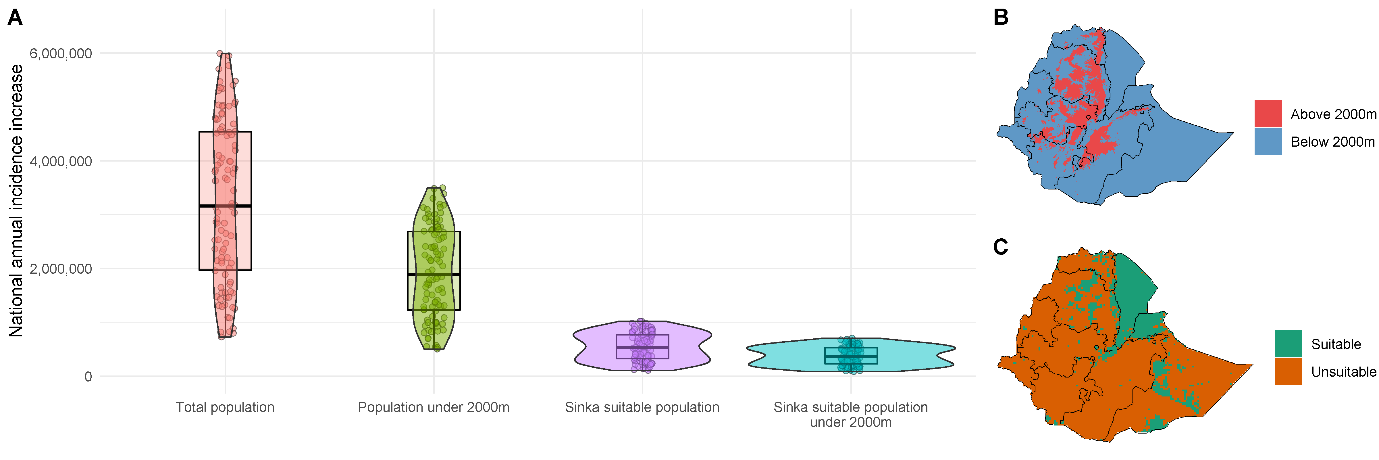

**Additional File Figure 5. Sensitivity analyses given uncertainty of the range of environments suitable for *An. stephensi* malaria transmission.** A) Comparison of different population denominators on the estimates of increases to malaria incidence, B) Ethiopian areas over/under 2000m altitude and C) Sinka et al., (2020) estimates of suitability of the environment to *An. stephensi* (Sinka suitable in A) normalised to a 0-1 scale, the cut-off assumed was 0.5. Coloured points in panel A, indicate the calculated annual incidence increase for individual LHC runs. Four different situations are considered; (i) – total population where everywhere in Ethiopia is suitable for *An. stephensi* establishment and malaria transmission is possible at altitudes above 2000m, (ii) *An. stephensi* establishment everywhere but malaria transmission only in areas beneath 2000m altitude, shown in panel B (iii) *An. stephensi* establishes in areas identified by Sinka et al. 2020 to be suitable, shown in panel C, but transmission possible in areas above 2000m, and (iv) areas restricted to those identified by Sinka et al and below 2000m in altitude (matching results shown in main text). Population denominators for the different groupings are shown in Additional File Table 3.

**Additional File Table 5. Populations and increase in annual clinical incidence from pre-*An. stephensi* era to after establishment for different scenarios.** Sinka suitable denotes areas where the environment has been predicted to be suitable to *An. stephensi* establishment by Sinka et al., (2020).

| Population type | Population size | Additional cases (95% CI) |
| --- | --- | --- |
| Total population | 114,139,000 | 3,163,000 (818,000 - 5,740,000) |
| Population under 2000m | 63,564,000 | 1,884,000 (542,000 - 3,348,000) |
| Sinka suitable population under 2000m | 12,410,000 | 368,000 (103,000 - 664,000) |
| Sinka suitable population | 18,757,000 | 531,000 (134,000 - 979,000) |

**Relationship of EIP and increases in prevalence**

Areas at higher altitude are predicted to have lower values of temperature dependent EIP (Additional File Figure 7). Following the establishment of *An. stephensi*, there is predicted to be minimal increase in malaria prevalence in areas with high EIPs, those at higher altitudes, compared to those at lower altitudes, and so lower EIPs. This is because the average time for a mosquito to acquire *Plasmodium* infection and then become infectious begins to eclipse the expected lifespan of the mosquito. In these areas even if *An. stephensi* were to establish, it is assumed to have a minimal impact on local malaria transmission.

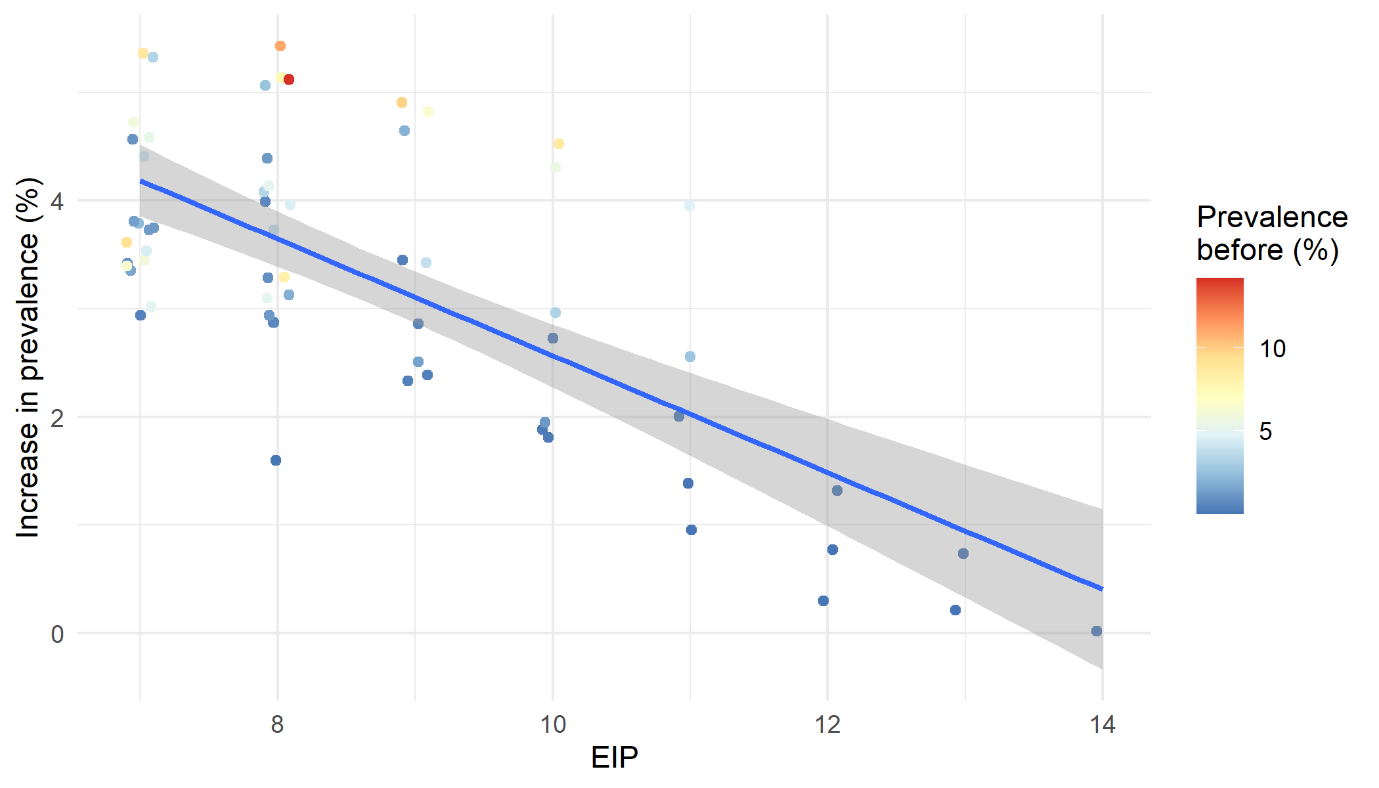

### **Additional File Figure 6**. **Relationship of absolute prevalence increase following *An. stephensi* introduction and estimated extrinsic incubation period (EIP) across Ethiopia.** Points are jittered on the x-axis to aid interpretation but are integer values in the data. Line represents a linear regression of the relationship of increase in prevalence and EIP. Colour of points refers to the prevalence in all ages pre-*An. stephensi* introduction.**4. Limitations and opportunities for data collection**

#### **Additional limitations to the modelling approach**

Initially mosquito invasion dynamics have been simplified, assuming invasion occurs equally in all locations modelled and that human-to-mosquito densities universally reach the same level as observed in Djibouti. This is unlikely to be the case but is currently the most parsimonious explanation in the absence of other information. Certain locations will be more or less suitable due to local ecological and anthropological conditions, as well as inter-species competition with other mosquito species. This will change both the presence/absence of the species but also their relative abundance. While not accounting for these directly, we have tested the presence/absence assumption by only predicting increases into areas that have been previously estimated to be suitable for *An. stephensi* following an early geostatistical analysis^[31]^ and in those regions under 2000m. While this makes a substantial difference to the overall cases, we still estimate an additional 368,000 (103,000 - 664,000) cases (compared to 3,163,000 (818,000 - 5,740,000) if the whole country is suitable). This different geographical spread will substantially affect the cost of scaling up interventions. Our current model ignores importation of disease through human movement, which could reduce the time between mosquito invasion and increases in disease. Additionally, *An. stephensi* is a highly competent vector for *P. vivax*, found throughout large parts of Ethiopia^[59]^. We have not considered malaria caused by this parasite, and so overall levels of malaria (independent of *Plasmodium* species) are likely to be higher than we have estimated. The model used to quantify the vector density required to explain malaria incidence in Djibouti, and subsequently predict the potential impact in Ethiopia, is a long standing and extensively tested model against real-world data^[19, 21-23, 28, 60]^ . This model was created and utilized in order to model *P. falciparum*, and as such has not been adequately constructed or parameterized for modelling *P. vivax* infection. Due to differences in the progression of disease (primarily relapsing in *P. vivax* malaria due to hypnozoites), predictions of vector density and subsequent impact will therefore be very different between *Plasmodium* species. Noting this, and the greater public health impact of *P. falciparum* infection, we have focused this body of work on *P. falciparum*, while acknowledging this is a limitation in establishing the true potential impact of *An. stephensi*.

The vector may additionally be capable of establishing outside of areas previously predicted to be suitable, and by including the effect of a temperature dependent extrinsic incubation period (EIP), we can partially account for the effect of altitude on malaria transmission. This is shown by many of the regions at higher altitudes, with longer EIPs, have lower increases in prevalence (Additional File Figure **6**). Furthermore, we are not assessing invasion dynamics or timelines, and instead have looked at the differences pre- and post- *An. stephensi* establishment. A more realistic increase in burden would be staggered as establishment will likely vary, which has been seen in *An. stephensi* primarily being detected in eastern and central Ethiopia, so far^[59]^. Finally, we have used a deterministic model structure. The invasion of *An. stephensi* and the subsequent rise of malaria cases is likely to be a highly stochastic process in space and time. This will make predictions of public health impact and the effect of vector control interventions highly uncertain. While mechanistic malaria models have long been used for predicting the impact of interventions in endemic settings^[19, 21, 22, 60, 61]^, this model has not been validated when considering invasion into an area, and so aspects such as the acquisition of immunity in low transmission scenarios should be treated with extreme caution.

This paper has identified a number of different hypotheses such as the speed in the rise of malaria cases will depend on pre-*An.stephensi* malaria endemicity. These hypotheses need to be verified by good quality local surveillance data and models adjusted accordingly.

#### **Future data collection to inform mathematical modelling.**

Improved understanding of the current entomological and epidemiological situation in regions where *An. stephensi* may invade will improve projections of its potential public health impact and how effective mitigation measures will be (Additional File Table 6. . This analysis has highlighted how the increase in malaria burden depends on current malaria endemicity, so more detailed knowledge of the heterogeneity in malaria prevalence and the existing use of vector control interventions in urban and peri-urban areas where the mosquito might invade will be key to understanding overall impact. Malaria burden is also heavily dependent on the abundance of the invading mosquito species and so an understanding of the carrying capacity of the species in the new environment (and how this varies between regions) will enable more tailored projections.

Uncertainty in vector bionomics and behaviours have necessitated several assumptions in this modelling framework. These unknowns, and the sampling structure designed to compensate for them, introduce substantial uncertainty into the results. While a level of uncertainty is expected, with further data on the vector and its role in transmission this can be substantially reduced. Some of the most important vector parameters identified by the model to influence the impact of *An. stephensi* invasion and its control are listed in Additional File Table 6 whilst results from a univariate sensitivity analyses showing how mosquito bionomics influence post-An. *stephensi* malaria incidence is given in Additional File Figure 7. We also list the important factors determining intervention effectiveness, many of which will depend on the level of effort deployed. An understanding of the price of these different levels of effort will allow further refinement of the cost-effectiveness analyses.

**Additional File Table 6. Key parameters influencing modelling results and the aspects of mathematical modelling they inform.**

| Parameter type | Parameter | What it informs |
| --- | --- | --- |
| Vector bionomic/ behaviour | Life-expectancy *of* *An. stephensi* in Africa | Disease endemicity and impact of interventions |
|  | Anthropophagy (human blood index) | Disease endemicity and impact of interventions |
|  | Endophily (mosquito resting behaviour) | Impact of IRS |
|  | Proportion of mosquito bites taken when people indoors | Impact of IRS and ITN |
|  | Proportion of mosquito bites taken when people in bed | Impact of IRS and ITN |
|  | Sporozoite rate in different vector species | Estimate of the relative significance of the invading mosquito population in relation to other local species. |
|  | Seasonality of *An. stephensi* abundance | Impact and requisite frequency of IRS |
| Intervention Efficacy | Level of pyrethroid resistance | Effectiveness of ITNs (both pyrethroid-only and pyrethroid-PBO nets) |
|  | Percentage of mosquito resting sites accessible to IRS campaigns. | Impact and requisite frequency of IRS |
|  | Durability of IRS in structures in the region (will vary between products) | Impact and requisite frequency of IRS |
|  | Reduction in emergence of adult mosquitoes due to larval source management | Impact of larval source management |

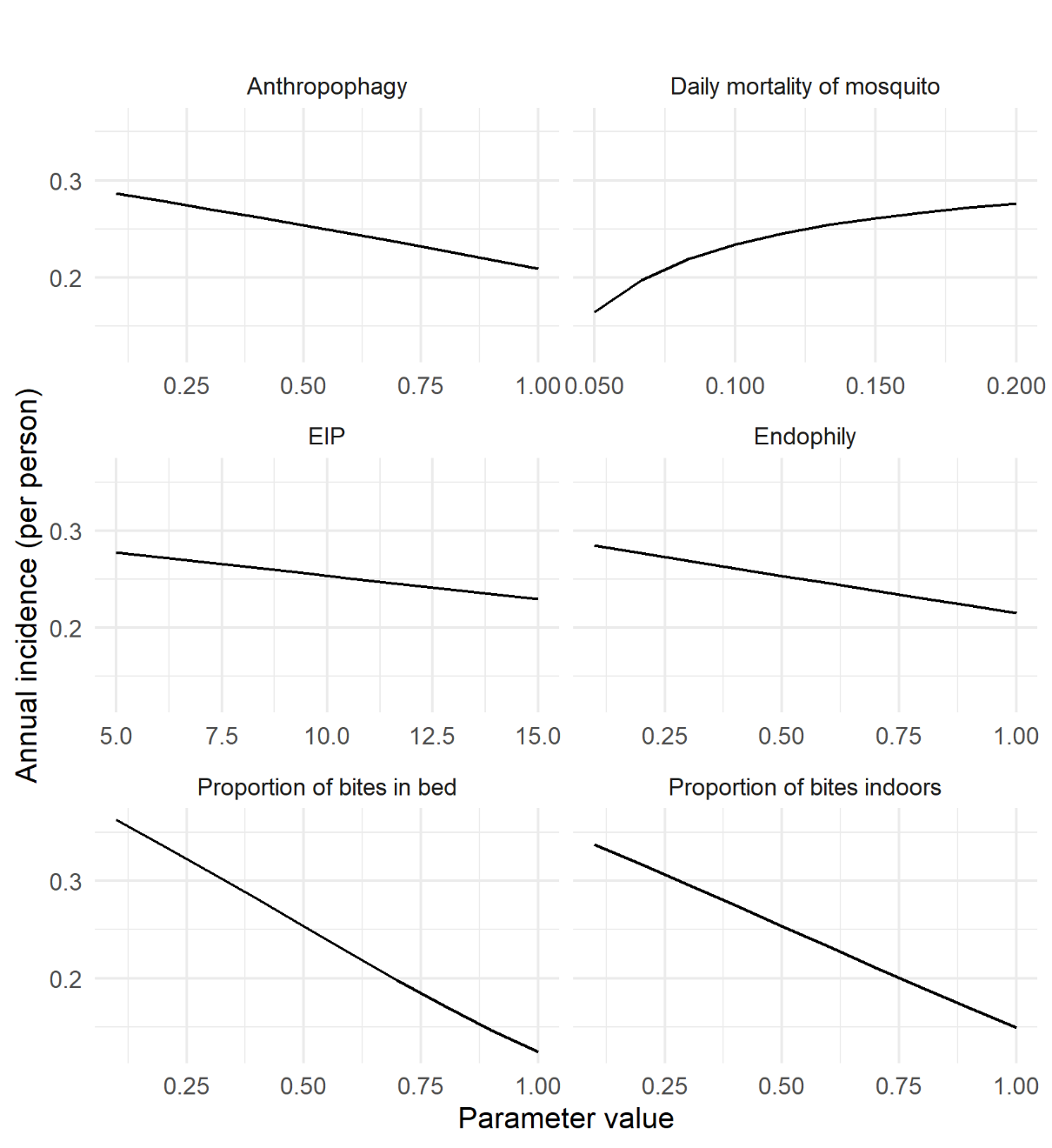

**Figure 7. Univariate sensitivity analysis for entomological parameters varied in the model.** Results are shown for model predictions of the number of clinical cases per person (per year) in a site with a pre-*An. stephensi* entomological inoculation rate (EIR) of 10 and following the introduction of 50% ITN and IRS coverage after mosquito invasion. EIP is measured in days, the other values are a proportion (0-1).
